# Remote Ischemic Conditioning Modulates Spinal Reflex Excitability in Healthy Young Adults: A Pilot Randomized Controlled Trial

**DOI:** 10.64898/2026.10.28.26362269

**Authors:** Shailesh S. Gardas, Sarah Baker, Maz J. Kurz, John Willson, Chia-Cheng Lin, Erzsebet Szatmari, Linda May, Swati M. Surkar

**Author notes:** **Registry** ClinicalTrials.gov, Registration No. NCT06860464.

## Abstract

**Background:** Remote ischemic conditioning (RIC), involving transient and sublethal bouts of ischemia and reperfusion, promotes neuroplasticity and motor learning. Although early clinical studies suggest supraspinal and muscular mechanisms underlying RIC, its influence on spinal reflex excitability – a key substrate for motor learning – remains poorly understood. Balance training alone is known to modulate spinal excitability, but whether RIC can enhance this training-induced adaptation has not been tested.

**Objective:** This prospective single-blind, pilot randomized controlled trial (RCT) explored the preliminary effects of pairing RIC with balance training on spinal reflex excitability and balance performance in healthy young adults and generated estimates of variability to inform a future definitive randomized trial.

**Methods:** Twenty-six participants aged between 18-40 years were randomized to RIC or sham groups at university-based research setting between March and August 2025. The intervention included five cycles of 5-minute cuff inflation (RIC: 200 mmHg; sham: 25 mmHg) and 5-minute deflation on the dominant thigh, followed by balance training on a stability platform (15 trials/day) for five consecutive days. Pre- and post-intervention assessments included Hoffmann’s reflex (H-reflex) parameters—H_max_ (peak H-reflex amplitude reflecting motor neuron recruitment via Ia afferent input), H/M ratio (H_max_ normalized to maximal M-wave amplitude), H_slp_/M_slp_ ratio (relative slope of the H-reflex and M-wave recruitment curves), and H-reflex threshold (minimum stimulus intensity eliciting a response)—used to index spinal motor neuron pool excitability, as well as balance performance, quantified as center time (the duration participants maintained a stability platform within a set horizontal threshold). Independent sample and Mann-Whitney U tests analyzed the differences in the change scores (RIC vs Sham) for H-reflex and balance outcomes respectively.

**Results:** The RIC group showed directionally greater reduction in the H_slp_/ M_slp_ ratio (p = 0.02) and greater improvement in balance performance (p = 0.02) compared to sham group. No significant between-group differences were observed for H_max_, H/M ratio, or H-reflex threshold; however, the direction of changes in Hmax was consistent with reduced spinal reflex excitability following RIC. These findings potentially suggest that RIC could augment the neuroplastic adaptations typically induced by balance training.

**Conclusion:** This pilot study provides preliminary evidence that RIC paired with balance training may influence selected spinal neurophysiological responses and improve balance performance in healthy adults. The potential convergence of neurophysiological and behavioral adaptations observed in this pilot study may suggest a role for RIC as a physiological primer for sensorimotor learning. Moreover, these findings establish a foundation for future large-scale trials incorporating cortical and presynaptic inhibitory measures to elucidate mechanisms and clinical applicability in neurological populations.

## Introduction

Ischemic conditioning (IC) describes an endogenous phenomenon in which brief, sublethal episodes of ischemia followed by reperfusion protect tissue from future subsequent severe ischemic injury (1). First demonstrated by Murry et al. in canine heart muscle, this concept was initially applied through direct ischemia to target organ (2). Remote ischemic conditioning (RIC) extends this principle where transient ischemia in a remote tissue, such as a limb, can confer protection from ischemia-reperfusion injury to distant organs including the brain (3,4). Clinically, RIC is a practical, non-invasive method delivered through repeated inflation and deflation of a blood pressure cuff on a limb (5).

Since its introduction in the late 1980s, RIC has demonstrated pleiotropic benefits extending well beyond its original application in cardioprotection. Recent evidence indicates its role in neuroprotection (6,7), muscle performance (8), and motor learning (9). Preclinical studies consistently show that RIC attenuates ischemic brain injury by reducing infarct size, limiting cerebral edema, and accelerating neurological recovery (10,11). In healthy individuals, even a single session of RIC has been associated with increased muscle force, delayed fatigue onset, and improved endurance and oxygen uptake in the upper and lower limb muscles (12–14). When combined with strength training, RIC augments muscle strength beyond those achieved with training alone (8). When paired with task-specific balance training, RIC also enhances dynamic balance performance and retention in healthy young adults (9). Clinical studies further demonstrate benefits in neurological populations: RIC improves walking capacity and muscle torque while reducing fatigability in stroke population (15,16), enhances walking performance in multiple sclerosis (17), reduces biomarkers of neuronal injury following traumatic brain injury (18), and increases probability of favorable recovery after stroke (19). Moreover, in ALS and spinal cord injury, RIC has also been shown to enhance motor evoked potentials indicating enhancement of cortical neuroplasticity (20).

Although the precise mechanisms mediating the effects of RIC remains incompletely defined, current evidence suggests a complex interplay of neural, metabolic, and humoral pathways (21,22). Within the CNS, RIC promotes neurogenesis, angiogenesis, and synaptic plasticity by modulating N-Methyl-D-Aspartate receptor activity, enhancing gamma-aminobutyric acid (GABA) synthesis, and upregulating brain-derived neurotrophic factor (BDNF)—mechanisms that overlap with motor learning and cortical plasticity (23,24). Peripherally, RIC augments muscle function by attenuating mitochondrial ATPase activity, lowering energy demands, and slowing ATP depletion and lactate production (25,26). In addition to these cortical and metabolic mechanisms, experimental evidence suggests that RIC engages peripheral sensory and autonomic pathways that converge on spinal cord (27–29), thereby implicating spinal networks as a potential substrate for its effects.

Spinal neural networks represent a critical substrate for motor skill acquisition and neuroplasticity (27). The Hoffmann reflex (H-reflex) provides a robust index of alpha motor neuron pool excitability and is widely used as a surrogate marker of spinal plasticity (30). Task-specific trainings distinctly modulate H-reflex excitability in healthy adults—strength (31,32) and endurance training (33) enhance excitability, whereas balance training attenuates it (34,35). In neurological populations, heightened H-reflex excitability is often associated with spasticity and impaired function (36,37), but locomotor-based interventions can normalize excitability and improve functional outcomes (38,39). These findings highlight the importance of spinal mechanisms in functional recovery and raise the possibility that RIC may modulate spinal reflex pathways to optimize postural stability – a question that remains largely unexplored.

Despite this plausibility, and growing evidence for RIC’s neuromodulatory effects (18,19,21,22), evidence for RIC-induced spinal modulations is scarce. Only one study to date has tested this hypothesis, reporting greater modulations in spinal reflex excitability as measured by H-reflex measures compared to sham conditioning (40). However, methodological differences–– including a short washout period and higher than the standard pressure used for sham condition (40) –– may limit the interpretation of those findings. Thus, more rigorous trials are needed to establish whether RIC influences spinal excitability in humans.

Therefore, the primary aim of this pilot randomized controlled trial (RCT) was to investigate whether RIC combined with balance training modulates spinal reflex excitability and enhances balance performance in healthy young adults. We examined whether changes in H-reflex parameters and balance performance showed directional patterns favoring RIC compared with sham conditioning, without undertaking confirmatory efficacy testing. By exploring potential spinal mechanisms of RIC-induced spinal neuroplasticity, this pilot trial provides preliminary evidence to inform future larger clinical trials in healthy adults and eventually translate into studies aimed at optimizing functional recovery in individuals with neurological impairments.

## Materials and methods

### Trial design

This study was a prospective, single-blind, pilot RCT. The protocol was approved by the Institutional Review Board of East Carolina University (UMCIRB 23-001021) in January 2025, and the trial was conducted in the university setting between March and August 2025. The authors confirm that all ongoing and related trials for this intervention is registered at clinicaltrials.gov (NCT06860464). All participants provided written informed consent and received financial compensation for their participation. The study was performed in accordance with the ethical standards of Declaration of Helsinki.

### Participants

Healthy young adults between 18-40 years were recruited via emails circulated through the listservs of various colleges at East Carolina University. The emails included an IRB-approved study flyer and a QR code linked to a REDCap screening survey to determine eligibility. Figure 1 illustrates the CONSORT diagram, detailing how participants progressed through the study, including withdrawals, and their inclusion in the analysis.

**Figure 1.** CONSORT diagram. A total of 110 participants were assessed for eligibility, and 30 participants were eventually randomized. Fifteen participants each were assigned to receive RIC and Sham conditioning groups. Per the CONSORT diagram, 26 participants were included in the final analysis (RIC: n = 13; sham: n = 13).

Inclusion criteria were: (1) age 18 to 40 years, (2) and intact cognitive-motor functions to actively participate. Exclusion criteria were: (1) a history of any neurological condition ( i.e. stroke, Alzheimer’s disease, Parkinson’s disease), diagnosed psychiatric conditions (such as attention deficit hyperactivity disorder, depression, or bipolar disorder), balance impairment, or vestibular disorder; (2) recent lower extremity injuries or surgeries within last three months which could compromise balance training; (3) any extremity, soft tissue, orthopedic, or vascular injury (i.e. peripheral vascular disease) which may contraindicate RIC; (4) a history of sleep apnea which could confound the effects of RIC; (5) any cognitive, sensory, or communication problem that might prevent completion the study; (6) moderate or vigorous intensity of exercises for more than 3 days/ week, which could confound the effects of RIC or alter spinal excitability; (7) current substance abuse or dependence; (8) current use of medications, such as selective serotonin reuptake inhibitors, that could decrease nervous system excitability and alter spinal excitability; (9) participation in previous RIC studies; and (10) an inability or unwillingness to travel for all study visits.

### Sample Size and Randomization

Established guidelines for pilot studies with continuous outcomes suggest a sample size of 10 - 20 participants per arm when anticipating a small to medium standardized effect size for a future main trial with 80% power. This range is consistent with recommendations from Julious, who suggests a minimum of 12 participants per arm, and Kieser and Wassmer, who recommend between 10 and 20 per arm (41). Accordingly, in this pilot study we recruited 15 participants per group to estimate the variability of the primary outcome measure. This pilot study was not designed to provide confirmatory evidence of differences between groups. Accordingly, the findings from this study were interpreted as descriptive and exploratory, with emphasis on outcome variability, estimated between group differences, and 95% confidence intervals to inform outcome selection and sample size planning for a future definitive trials (42).

Participants were randomly assigned (allocation ratio 1:1) by the PI to either the RIC or sham group using the Randomization.com (RR) app. Allocation was concealed using sequentially numbered, opaque envelopes prepared by a researcher not involved in participant enrollment or assessment. Only the participants were blinded to the group assignment, while the investigators delivering the intervention were aware of the allocation.

### Experimental procedures

The graphical schematic of experimental procedures is shown in Figure **2**. The study involved a total of seven consecutive visits. During pre-intervention, we collected demographic information, including height, weight, lower limb dominance, health and exercise history, calf girth (dominant leg), and conducted baseline (pre-testing) assessments of H-reflex and balance performance. Visits 2 to 6 consisted of RIC combined with balance training. Post-intervention involved post-testing of H-reflex and balance performance.

**Figure 2.** Graphical schematics of study protocol.

### Interventions

#### Remote Ischemic Conditioning

The intervention was delivered by the research team member. Before beginning RIC or sham conditioning, we recorded resting heart rate, blood pressure (BP), and oxygen saturation. Conditioning was performed in the supine position through cyclic inflation and deflation of a pressure cuff on the dominant mid-thigh using a using a Hokanson rapid cuff inflator (Hokanson, Inc.) (Figure 3A). Both the RIC and sham conditioning groups received five cycles of 5-minute cuff inflation followed by 5 minutes of deflation, resulting in a total conditioning time of 45 minutes. The conditioning pressure in the RIC group was maintained at 200 mm Hg, while in the sham group, it was maintained at 25 mm Hg, which produces the sensation of cuff inflation without inducing arterial occlusion (4,16). All participants received conditioning during visits 2 to 6. Throughout the intervention, the experimenter continuously monitored for the presence or absence of ischemia using a pulse oximeter placed on big toe of the conditioned lower limb and by visually inspecting the discoloration of the legs. A pulse and oxygen saturation reading of “0” on the oximeter, along with a pale, dusky appearance of the leg, confirmed the presence of ischemia in the RIC group. In contrast, stable pulse and oxygen saturation values equivalent to baseline, along with unchanged color of the limb, confirmed the absence of ischemia in the sham group. In the RIC group, if a pulse or oxygen saturation reading appeared on the oximeter during an inflation cycle, the interventionist increased the cuff pressure until ischemia was confirmed. The inflation duration was adjusted to ensure a complete 5-minute ischemic period. In the sham group, if oxygen saturation or pulse dropped below baseline, the cuff pressure was reduced until baseline values were restored and the limb showed no visible signs of ischemia.

**Figure 3.** A. Remote Ischemic and Sham Conditioning Protocol – Participant in supine position with a Hokanson cuff placed on the dominant thigh. Conditioning pressure was set at 200 mmHg for the RIC group and 25 mmHg for Sham. A pulse oximeter on the toe of the conditioned limb was used to confirm presence or absence of ischemia and monitor vitals (O₂ saturation and heart rate) for safety. B. Dynamic Balance – Participant performing a dynamic balance task on the Lafayette stability platform, aiming to maintain the platform within a ±5° threshold.

Cardiovascular stability was ensured by monitoring the systolic and diastolic BP, oxygen saturation, and heart rate on the arm before, during, and after each session. Pain due to conditioning was also monitored. Conditioning was terminated for both groups if any of the following occurred: (1) systolic BP <85 or >140 mm Hg; (2) diastolic BP <50 or >90 mm Hg; (3) heart rate <60 or >115 bpm; (4) oxygen saturation <90%; or (5) pain exceeded 6 on a 10-point Likert pain scale.

#### Dynamic Balance

As per established protocol (9,43), we used a stability platform (Lafayette Instrument, Model 16030L; Lafayette, IN) for testing and training dynamic balance (Figure 3B). It’s an ecologically valid motor learning task that engages multiple systems, including the vestibular, visual, motor, somatosensory, and cognitive systems (44). Successful performance on the stability platform requires participants to anticipate and coordinate their lower limbs with their trunk and upper limbs in response to self-induced perturbations.

Participants were instructed to stand on the stability platform with their feet facing forward and were secured by an overhead harness for safety. The goal of the task was to keep the platform level within a ±5° for as long as possible during 30-second trial. Prior to testing, two familiarization trials were provided. Each training session consisted of 3 blocks of 5 trials (15 trials/ visit). Each trial lasted for 30 seconds, followed by 30 seconds of rest and between each block, with a 2-minute rest period between blocks. Performance was quantified by measuring the cumulative amount of time that a participant maintained the platform within ±5° of horizontal during each trial. Participants performed a total of 75 training trials during visits 2 to 6 (15 trials per day) immediately (within 5 minutes) after conditioning. During pre- and post-testing, participants performed 5 trials each. The performance was automatically recorded by the device. Use of the handrail was permitted for familiarization trials and feet positioning; however, no upper extremity support was allowed during a trial. After each trial, participants received knowledge of their performance–– the number of seconds they successfully maintained the platform in balance. Balance training was performed immediately after the conditioning session.

### Spinal (Hoffmann’s) Reflex Excitability Testing

Hoffmann’s (H-reflex) testing was conducted to assess spinal motor neuron pool excitability. All participants were positioned in the prone with arms by their sides, head turned to one side, eyes closed, and a pillow placed under the ankles to allow the feet to hang off the edge of the bed. This position minimizing voluntary muscle activity thereby optimizing acquisition of maximum H-reflex amplitude with minimum variability (30,45). To minimize confounding influences on spinal reflex excitability, participants were instructed to abstain from caffeine intake prior to testing, and all H-reflex assessments were conducted at approximately the same time of day for each participant across pre- and post-intervention visits.

Surface electromyography (EMG) was recorded from the soleus (Sol) muscle of the dominant limb using self-adhesive Ag/AgCl electrodes, with 2 cm interelectrode distance aligned parallel to the muscle fibers over the belly of soleus muscle. Prior to electrode application, the skin over the muscle was cleaned with 70% isopropyl alcohol prep pads to remove debris and reduce skin-electrode impedance. Electrodes were positioned lateral to the midline of the calf and 4 cm below the inferior margin of the gastrocnemius muscle, in accordance with surface EMG for non-invasive assessment of muscles (SENIAM) guidelines (Figure 3A). Electrode placement was documented with photographs and anatomical measurements to ensure consistent positioning across testing sessions (Figure 4A) (46,47).

**Figure 4:** H-reflex testing. A. Position of the participant (prone) with electrode placement for stimulating (on the popliteal fossa proximal and distal to knee joint crease) and recording (four centimeters distal inferior margin of gastrocnemius muscle bulk and lateral to midline) electrodes. B. Constant-current stimulator (Digitimer DS8R) used to deliver single rectangular pulses (1-ms pulse duration, 0.1 Hz). C. Representative recruitment curve showing the relationship between stimulus intensity (mA) and response amplitude (mV). H_max_ and M_max_ are determined using RC and used to calculate H/ M ratio. Bold (green lines) are H slope and M slope determined by using linear regression function in Microsoft excel.

Participants underwent lower extremity H-reflex testing using a constant-current stimulator (Digitimer DS8R, Ft. Lauderdale, FL; Figure 4B). Percutaneous nerve stimulation was performed to elicit H-reflexes in the dominant Sol muscle by stimulating the tibial nerve with surface electrodes positioned at the midline of the popliteal fossa, with the cathode proximal and the anode distal to the knee joint crease (48). To account for changes at the skin–electrode surface, maximal M-waves (M_max_) were elicited in the Sol muscle and utilized for normalization of the H-wave responses.

### Data Processing – Spinal Reflex Excitability

After testing, the EMG signals were amplified, band-pass filtered (20–500 Hz), and sampled at 1926 Hz for both real-time and offline analysis using a custom MATLAB program. Stimulus-response recruitment curve (Figure 4C) was generated by delivering single rectangular pulses (1-ms pulse duration, 0.1 Hz) beginning at 0 mA and increasing in 2 mA increments every 10 seconds until the participant’s tolerance was reached, and a maximal muscle (M) response was achieved. A stimulation frequency of 0.1 Hz (inter-stimulus interval of 10 seconds) was used to prevent the influence of inhibitory mechanisms on H-reflex amplitude. M max was defined, through visual inspection, as the point beyond which the M-wave amplitude no longer increased with further increases in current intensity. The maximal H-reflex (H_max_) and M max were used to calculate the H_max_/ M_max_ ratio (H/M ratio) (46).

Slopes of the ascending limbs (Figure 4C) of the H-reflex (H_slp_) and M-wave (M_slp_) recruitment curves were calculated to reduce inter- and intra-subject variability and to minimize the sensitivity of H-reflex facilitation and inhibition near H_max_ (49–53). The linear portion of each recruitment curve was defined as the segment between 25% and 80% of the maximal response. Within this range, slopes were determined using the linear regression function in Microsoft Excel (=SLOPE (y_range, x_range)), where y_range represented the response amplitudes of H-reflex and M-wave, and x_range represented their corresponding stimulus intensities. After determining the H_slp_ and M_slp_, the ratio of H_slp_/ M_slp_ was calculated to provide a standardized measure of motoneuron excitability. All the participants underwent H-reflex testing during visits 1 and 7 (30,40).

### Outcome measures

#### Primary H-reflex outcomes

1. *Maximal H-reflex amplitude (H_max_, mV):* H_max_ was defined as the peak-to-peak amplitude of the maximal H-reflex, reflecting the proportion of alpha MN recruited through monosynaptic Ia afferent input. An increase in H_max_ following training denoted increased H-reflex excitability and vice versa ^[40, 48]^. Hmax is a well-established measure of spinal reflex excitability with good between-session reliability (ICC = 0.88) (54).
2. *H/M ratio:* The H/M ratio is defined as the ratio of maximal H-reflex and maximal M-wave amplitudes – both (H_max_ and M_max_) are determined from the recruitment curve procedure. This is a widely used normalization method that minimizes peripheral influences such as electrode placement and muscle fiber conduction, providing a reliable estimate of motoneuron pool excitability. Excellent between-session reliability has been reported for the H/M ratio (ICC = 0.98), supporting its use as a reproducible measure of spinal reflex excitability (54). An increase in H/M ratio following training denotes increased H-reflex excitability and vice versa (40,47).
3. *Ratio of H-slope by M-slope (*H_slp_/ M_slp_): H_slp_ and M_slp_ is defined as the slope of H-reflex and M-wave. An increase in this ratio indicates greater spinal reflex excitability, whereas a decrease reflects reduced excitability. Slope-based recruitment curve analysis has demonstrated good test–retest reliability, with an ICC of 0.79 reported for regression slope measures, supporting its use as a reproducible outcome measure of spinal excitability (50).

#### Secondary H-reflex outcome

*4. H reflex threshold (milliamperes)*: It was defined as the minimum intensity of stimulus required to elicit the H-reflex. A decrease in H-reflex threshold following training denotes increased H-reflex excitability and vice versa (55).

#### Primary Balance outcomes

*5. Balance performance (center time in seconds):* Performance on the dynamic balance task was defined as the average amount of time participants were able to maintain the platform within ±5° of horizontal during the five trials conducted in the pre- and post-testing visits. Time-based balance measures obtained using the Lafayette Stability Platform have demonstrated high test–retest reliability (ICC = 0.74–0.76) and good within-session reliability (ICC = 0.71–0.86), supporting their reproducibility for assessing balance performance (56).

### Statistical Analysis

Data were analyzed using IBM SPSS Statistics Version 28.0.0 (Armonk, NY, USA). Descriptive statistics are presented as mean ± standard deviation (SD) for continuous variables. The distribution of continuous variables H-reflex (H_max_, H/M ratio, H_slp_/ M_slp_ ratio, and H-threshold) and balance performance (center time) variables were assessed using the Shapiro– Wilk test together with visual inspection of histograms and normal Q–Q plots. Change scores (Post – Pre) were calculated for each outcome to represent modulation in spinal reflex excitability and improvement in balance performance following the intervention. Between-group differences in change scores (RIC vs. Sham) were analyzed using independent samples t-tests for all normally distributed variables. When the normality assumption was violated, nonparametric Mann–Whitney U tests were performed (H/M ratio, H-threshold, and H_slp_/ M_slp_ ratio). Statistical significance was set at p < 0.05.

This study was designed as a pilot randomized controlled trial; hence, all inferential analyses were considered exploratory and were intended to provide preliminary estimates of intervention effects to inform future adequately powered trials. Therefore, emphasis was placed on confidence intervals rather than hypothesis testing. P-values are reported as exploratory measures and should not be interpreted as confirmatory evidence of intervention efficacy.

## Results

Twenty-six healthy adults, aged 18-40 years (RIC/ Sham, mean ± SD: 27 ± 5.3, 27.1 ± 5.9 years) participated in this study. Table 1 summarizes participant demographic characteristics included in the study.

**Table 1.** Baseline demographic details of the study participants.

| Characteristics | Participants |  | p - value |
| --- | --- | --- | --- |
|  | RIC (n=13) | Sham (n=13) |  |
| Sex, n (%) |  |  | 0.06 |
| Male | 8 (61.5%) | 3 (23.1%) |  |
| Females | 5 (38.5%) | 10 (76.9%) |  |
| Age, mean ± SD | $27 \pm 5.3$ | $27.1 \pm 5.9$ | 0.91 |
| Dominance, n (%) |  |  | 0.70 |
| Left | 2 (15.4%) | 2 (15.4%) |  |
| Right | 11 (84.6%) | 11 (84.6%) |  |
| Height (meters), mean ± SD | 1.71 ± 0.1 | 1.71 ± 0.1 | 0.51 |
| Weight (kgs), mean ± SD | 66.9 ± 10.3 | 81.5 ± 21.4 | 0.03* |
| BMI (kgs/ m<sup>2</sup>), mean ± SD | 22.88 ± 3.3 | 28.39 ± 7.3 | 0.01* |
| Calf girth (cms), mean ± SD | 34.76 ± 2.3 | 37.85 ± 4.9 | 0.07 |

\*denotes significant p value < 0.05. †Chi-square test for categorical variables (sex, dominance); ‡Independent samples t-test for continuous variables (age, height, weight, BMI, calf girth).

**Table 2:** Descriptive statistics of between-group comparison of neurophysiological and balance outcome measures for RIC and Sham groups.

| Outcome Variable | RIC Mean ± SD or Median [IQR] | Sham Mean ± SD or Median [IQR] | Statistic | p -value | 95% CI |
| --- | --- | --- | --- | --- | --- |
| $H_{\max}$ (mV)<sup>#</sup> | $-1.06 \pm 1.70$ | $-0.19 \pm 0.79$ | $t = -1.68$ | 0.11 | $[-1.95, 0.]$ |
| H/M Ratio<sup>†</sup> | $-0.47 [-10.38, 4.83]$ | $-0.53 [-6.49, 2.91]$ | $U = 78, Z = -0.33$ | 0.76 | — |
| $H_{\text{slp}}/M_{\text{slp}}$ Ratio<sup>†</sup> | $-2.97 [-6.59, -0.08]$ | $0.06 [-0.16, 0.53]$ | $U = 37, Z = -2.436$ | 0.016 * | — |
| H-Reflex Threshold (mA)<sup>†</sup> | $0.00 [0.00, 2.00]$ | $0.00 [0.00, 0.00]$ | $U = 77, Z = 0.39$ | 0.7 | — |
| Balance performance (s)<sup>#</sup> | 11.71 ± 2.84 | 9.2 ± 2.41 | t = 2.43 | 0.023 * | [0.38, 4.65] |

RIC = remote ischemic conditioning group; Sham = sham conditioning group. Data are presented as mean ± SD for normally distributed variables (analyzed with independent samples t-test denoted as #) or median [IQR] for non-normally distributed variables (analyzed with Mann-Whitney U test denoted as †). CI = confidence interval for the mean difference (t-test only). \* p < .05

### Comparison of Change Scores in H-reflex Measure - Spinal Reflex Modulation

1. *H_max_:* There was no significant difference observed between the RIC and Sham groups (*t* (25) = –1.68, *p* = 0.11, figure 5a). The RIC group demonstrated a mean reduction in Hmax of −1.06 ± 1.7 mV (95% CI [-2.089, -0.035]), compared with −0.19 ± 0.79 mV (95% CI [-0.663, 0.289]) in the Sham group, yielding a mean difference of −0.88 mV (95% CI [−1.861, 0.207]). Although the between-group difference was not statistically significant, the RIC group demonstrated a greater reduction in H_max_ than the Sham group, suggesting a directional trend toward reduced spinal reflex excitability.
2. *H/M ratio:* There was no significant difference between the RIC and Sham groups for the change in H/M ratio (*U = 78, Z = −0.33*, *p* = 0.76, figure 5b). The median changes in RIC group was −0.4 (IQR: −10.38 to 4.83) and the Sham group was −0.54 (IQR: −6.49 to 2.91), indicating negligible between-group difference in motoneuronal excitability.
3. *H_slp_/ M_slp_ ratio:* A nominal significant difference was observed for the change scores of H_slp_/ M_slp_ ratio, with the RIC group showing directionally greater reduction compared to Sham (*U = 37, Z = −2.44*, *p* = 0.02, figure 5d). The RIC group showed a median reduction of −2.97 (IQR: −6.59 to −0.08), compared with a median change of 0.06 (IQR: −0.16 to 0.53) in the Sham group. This may suggest that RIC may potentially modulate spinal reflex gain more effectively than sham conditioning.
4. *H-reflex threshold:* There was no significant difference between the RIC and Sham groups for H-reflex threshold (*U = 77, Z = 0.385*, *p* = 0.69, figure 5d). Both groups showed median change scores of 0.00 mA (RIC IQR: 0.0 to 2.0; Sham IQR: 0.0 to 0.0), indicating comparable shifts in recruitment threshold following both interventions.

### Comparison of Change Scores in Center Time – Balance Performance

A significant between-group difference (Figure 6) was observed for center time, a behavioral measure of postural control (*t* (25) = 2.43, *p* = 0.02, figure 6). The RIC group demonstrated a mean improvement in center time of 11.55 ± 2.8 s (95% CI [10.00, 13.42]) compared with 7.76 ± 4.20 s (95% CI [7.74, 10.66]) in the Sham group, yielding a mean difference of 3.8s (95% CI [0.99, 6.61]). Participants in the RIC group demonstrated a greater increase in center time compared to the Sham group, indicating greater improvement in balance performance following RIC.

**Figure 5.** Between-group differences in neurophysiological measures following RIC and Sham interventions. Bar graphs represent mean (± SE) change scores for a) H-reflex amplitude (H_max_), and box plots show median, interquartile range, and range for change scores for b) H/M ratio, c) H_slp_/ M_slp_ ratio and d) H-threshold. The RIC group demonstrated trends of greater reduction in Hmax and H_slp_/ M_slp_ ratio than the Sham group, whereas H/M ratio and H-reflex threshold showed comparable changes between groups, potentially indicating that RIC may have modulated spinal reflex more effectively than Sham.

**Figure 6.** Between-group differences in balance performance measures following RIC and Sham interventions. Bar graphs represent mean (± SD) change scores for center time. Independent *t*-tests revealed that the RIC group demonstrated a greater increase in center time than the Sham group indicating greater improvement in postural stability. This finding in conjunction with H-reflex changes could potentially support that RIC improves balance performance in healthy young adults due to the underlying spinal neurophysiological adaptations, however, these are only exploratory estimates informing future adequately powered studies.

Collectively, these results suggest that RIC did not produce statistically significant group-level changes in most measures of spinal reflex excitability (H_max_, H/M ratio, H-threshold) with the exception of H_slp_/ M_slp_ ratio, which showed a nominally significant greater reduction in the RIC group (p = 0.02). Although no significant between-group differences were observed for the remaining H-reflex measures, the direction of change was generally consistent with reduced spinal reflex excitability following RIC compared with sham. In addition, the significant enhancement in center time suggests that RIC may influence sensorimotor integration and postural stability, potentially through modulation of spinal-level neural mechanisms. Given the exploratory nature and limited sample size of this pilot study, these findings should be interpreted cautiously and used to inform the study design and sample size estimation of future adequately powered trials.

## Discussion

This pilot RCT provides exploratory findings suggesting that RIC, when combined with balance training, may influence spinal reflex modulation and balance performance in healthy adults. Compared with sham conditioning, the RIC group demonstrated a greater reduction in H-reflex outcomes and greater improvement in balance performance. The parallel direction of the neurophysiological and behavioral findings is consistent with the possibility that RIC may act as a neuromodulatory primer that supports adaptations associated with sensorimotor learning. However, given the exploratory design, small sample, and multiple outcomes examined, these findings should be interpreted as hypothesis-generating and require confirmation in an adequately powered trial.

Specifically, participants receiving RIC demonstrated a greater reduction in recruitment slope (H_slp_/ M_slp_ ratio) and directionally greater changes (reduction) in other H-reflex indices (H/M ratio and H_max_ amplitude) compared with sham conditioning. These neurophysiological differences were accompanied by improvements in dynamic balance performance, with the magnitude of change in the RIC group exceeding that observed in the sham group. However, substantial variability was observed across H-reflex measures, which may partly reflect baseline imbalances in sex distribution and BMI, both of which can influence spinal excitability. Given the pilot design and limited statistical power, these findings should be interpreted cautiously. Nevertheless, the parallel direction of changes in spinal reflex measures and balance performance suggests that RIC may influence neurophysiological adaptations associated with balance training and warrants further investigation in adequately powered trials. The directional trends observed in this pilot RCT was supported by prior work from Quadrado et al., who reported larger reductions in Hmax together with greater balance improvements following RIC compared with sham conditioning (40).

### RIC and Modulation of Spinal Reflex Excitability

Evidence shows that balance training alone is known to reduce spinal reflex excitability, reflecting a functional shift toward greater supraspinal contribution to postural control and reduced reliance on reflex-mediated responses (34,57–59). Consistent with this literature, both groups in the present study exhibited reductions in H-reflex measures following training. However, the larger directional changes observed in the RIC group—decreases in H_max_, the H/M ratio, and the H_slp_/ M_slp_ ratio of approximately 10%, 6%, and 40%, respectively, compared with minimal (1%) changes in the sham group— potentially indicating that RIC may have augmented these adaptations, although the present data do not allow definitive conclusions regarding the underlying mechanisms.

Previous work has demonstrated that balance training can increase cortical excitability and induce structural adaptations within motor-related brain regions, supporting a redistribution of control from spinal to supraspinal levels (60,61). Studies combining cortical and spinal measures further indicate that reduced spinal reflex excitability can coexist with enhanced corticospinal drive during postural tasks (62). Within this framework, RIC could potentially influence descending regulation of spinal circuitry and thereby modify training-related adaptations. This interpretation is biologically plausible given preclinical evidence that RIC engages systemic pathways associated with neural plasticity, including neurotrophic signaling and modulation of inhibitory neurotransmission (63,64), as well as early human evidence of changes in cortical excitability following RIC (17,18,65). However, cortical excitability and descending inhibitory mechanisms were not directly measured in the present study; therefore, a supraspinal contribution remains speculative.

RIC may also influence spinal excitability through peripheral afferent mechanisms. Endogenous mediators released during ischemic conditioning, including adenosine and bradykinin, have been proposed to alter group III and IV afferent signaling (29,66), which could indirectly influence motoneuronal excitability and reflex regulation. Such effects may interact with the reciprocal inhibitory and sensorimotor processes engaged during balance training (34,60,61). Nevertheless, because we did not directly assess afferent activity, presynaptic inhibition, corticospinal excitability, or muscle activation patterns, the physiological origin of the observed H-reflex changes cannot be determined. Future studies integrating cortical, spinal, and peripheral measures will be necessary to determine whether RIC-related modulation arises primarily from supraspinal influences, spinal circuit adaptations, peripheral mechanisms, or their interaction.

### RIC Enhances Dynamic Balance when Paired with Balance Training

The trends in neurophysiological adaptations in the RIC group were accompanied by greater improvements in dynamic balance performance. Overall, participants receiving RIC improved balance performance by 97% from baseline, compared with 70% in the sham group. These findings are consistent with prior work demonstrating that RIC combined with balance training can enhance balance performance and retention in healthy young and older adults (9,43). The relatively young age of participants in the present study likely contributed to improvements in both groups, yet the larger gains in RIC group could signal towards the priming potential of RIC in healthy populations.

The parallel direction of changes in spinal reflex measures and balance performance may also suggest a relationship between neurophysiological modulation and behavioral adaptation. One possibility is that RIC influences the interaction between descending motor control and spinal reflex regulation, thereby facilitating the adaptive suppression of reflex excitability required for dynamic postural control. RIC could also influence presynaptic inhibitory mechanisms that regulate Ia afferent input to the motoneuron pool. However, because presynaptic inhibition and corticospinal excitability were not directly assessed, the present study cannot determine whether the observed behavioral changes were mediated by spinal, supraspinal, or interacting mechanisms (67,68).

Taken together, the concurrent directional changes in H-reflex measures and dynamic balance performance provide preliminary support for further investigation of RIC as an adjunct to sensorimotor training. Future adequately powered studies incorporating cortical excitability, presynaptic inhibition, muscle activation, and spinal reflex measures are needed to determine whether these neurophysiological adaptations are mechanistically related to improvements in balance and to establish the potential relevance of RIC for neurological rehabilitation.

### Limitations and future directions

We acknowledge a few limitations in our study and propose directions for future research. First, H-reflex measurements were obtained at rest to minimize task-related variability, yet reflex modulation during active postural tasks may be more functionally relevant (35,60,69). Future studies should assess spinal excitability during standing or dynamic balance conditions. Second, group differences in sex and BMI may have influenced the neuromuscular responses and reflex modulation, as both factors are independently associated with altered H-reflex excitability (70,71). Larger, stratified trials are needed to address these potential confounders and establish reproducibility. Larger, stratified trials are needed to address these potential confounders and establish reproducibility. Third, PSI and corticospinal excitability were not directly measured, limiting mechanistic inference (37,40). Incorporation of transcranial magnetic stimulation and refined reflex paradigms will be important in future work. Finally, our sample size restricts generalizability and underscore the exploratory nature of these findings. In addition, formal tests of normality have limited power in small samples; therefore, the Shapiro–Wilk test was interpreted alongside graphical assessment of the data, and parametric analyses were selected based on the overall distributional characteristics and the robustness of the t-test to moderate departures from normality.

## Conclusions

The findings from this exploratory pilot study suggest that combining RIC with balance training is associated with directionally greater modulation of spinal excitability and enhancement of balance performance compared to sham conditioning. These preliminary observations indicate that RIC has the potential to influence neurophysiological and functional responses to balance training, supporting further investigation in adequately powered trials. Future studies in clinical populations with impaired postural control will be essential to determine whether RIC has translational value as an adjunctive strategy in neurorehabilitation.

## Data Availability

All data produced in the present study are available upon reasonable request to the authors

## Acknowledgements

We thank the participants, undergraduate & graduate research assistants for their contributions to this project.

## Author Contributions

Conceptualization: Swati M. Surkar, Shailesh S. Gardas, Maz J. Kurz

Data curation: Shailesh S. Gardas

Formal analysis: Shailesh S. Gardas, Swati M. Surkar.

Funding acquisition: Shailesh S. Gardas

Investigation: Shailesh S. Gardas, Swati M. Surkar

Methodology: Shailesh S. Gardas, Sarah Baker, Swati M. Surkar

Project administration: Swati M. Surkar

Resources: Swati M. Surkar.

Software: Shailesh S. Gardas, Sarah Baker, Max J. Kurz, Swati M. Surkar

Supervision: Swati M. Surkar, Maz J. Kurz, John Willson, Chia-Cheng Lin, Erzsebet Szatmari, Linda May

Validation: Swati M. Surkar

Visualization: Shailesh S. Gardas

Writing – original draft: Shailesh S. Gardas, Swati M. Surkar

Writing – review & editing: Swati M. Surkar, Maz J. Kurz, John Willson, Chia-Cheng Lin, Erzsebet Szatmari, and Linda May

## Data Availability Statement

All relevant data are within the paper and its Supporting Information files.

---

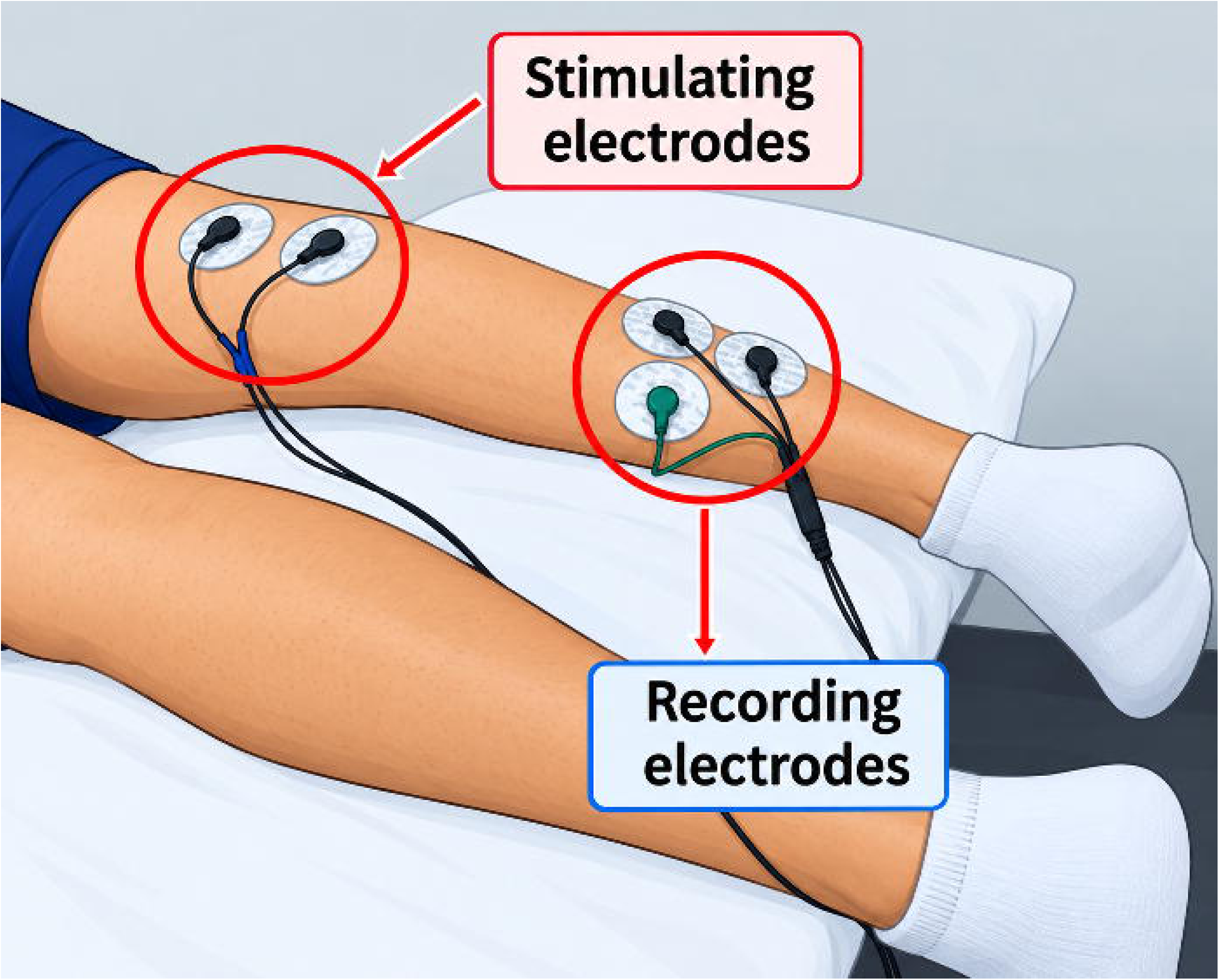

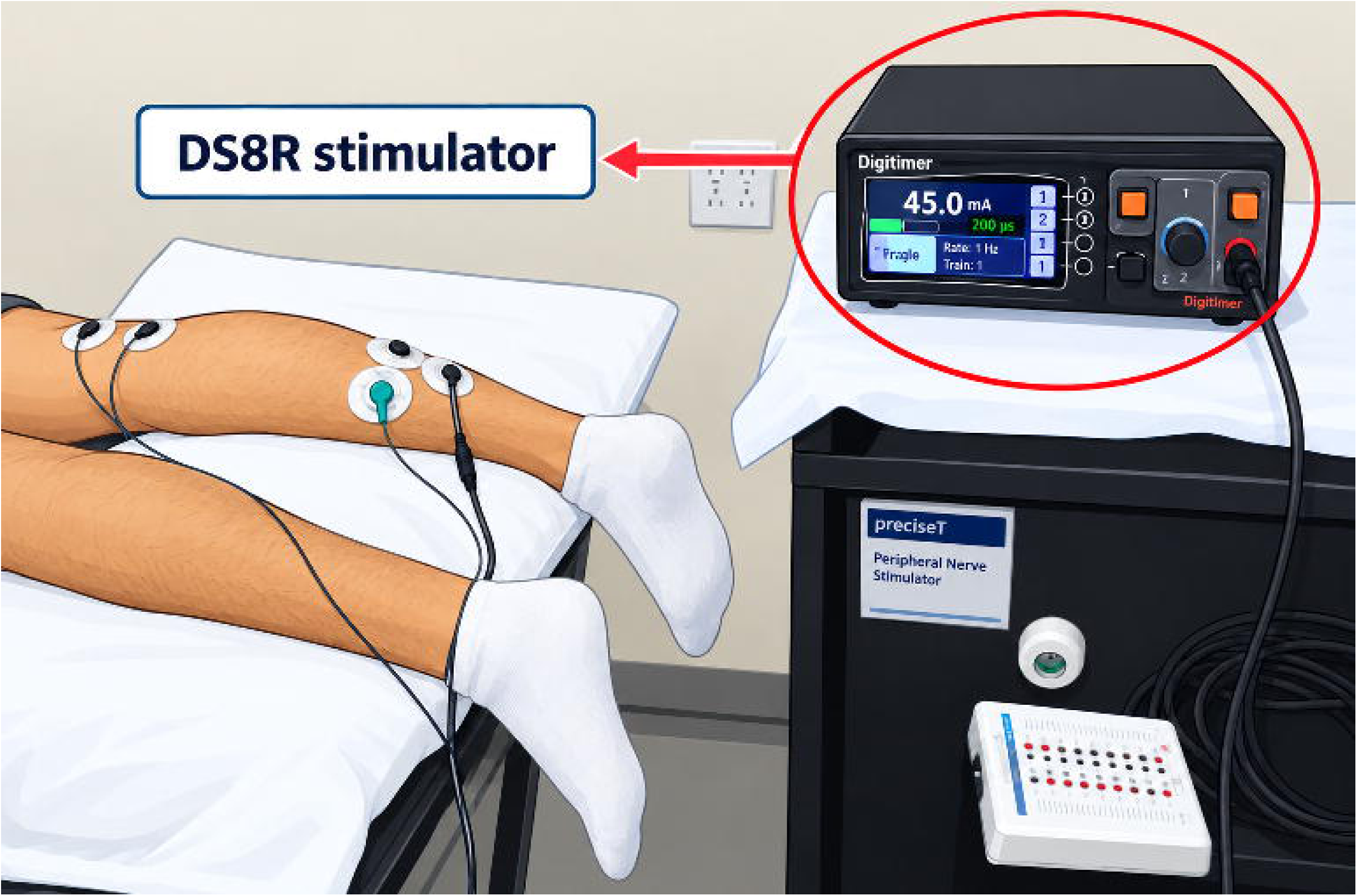

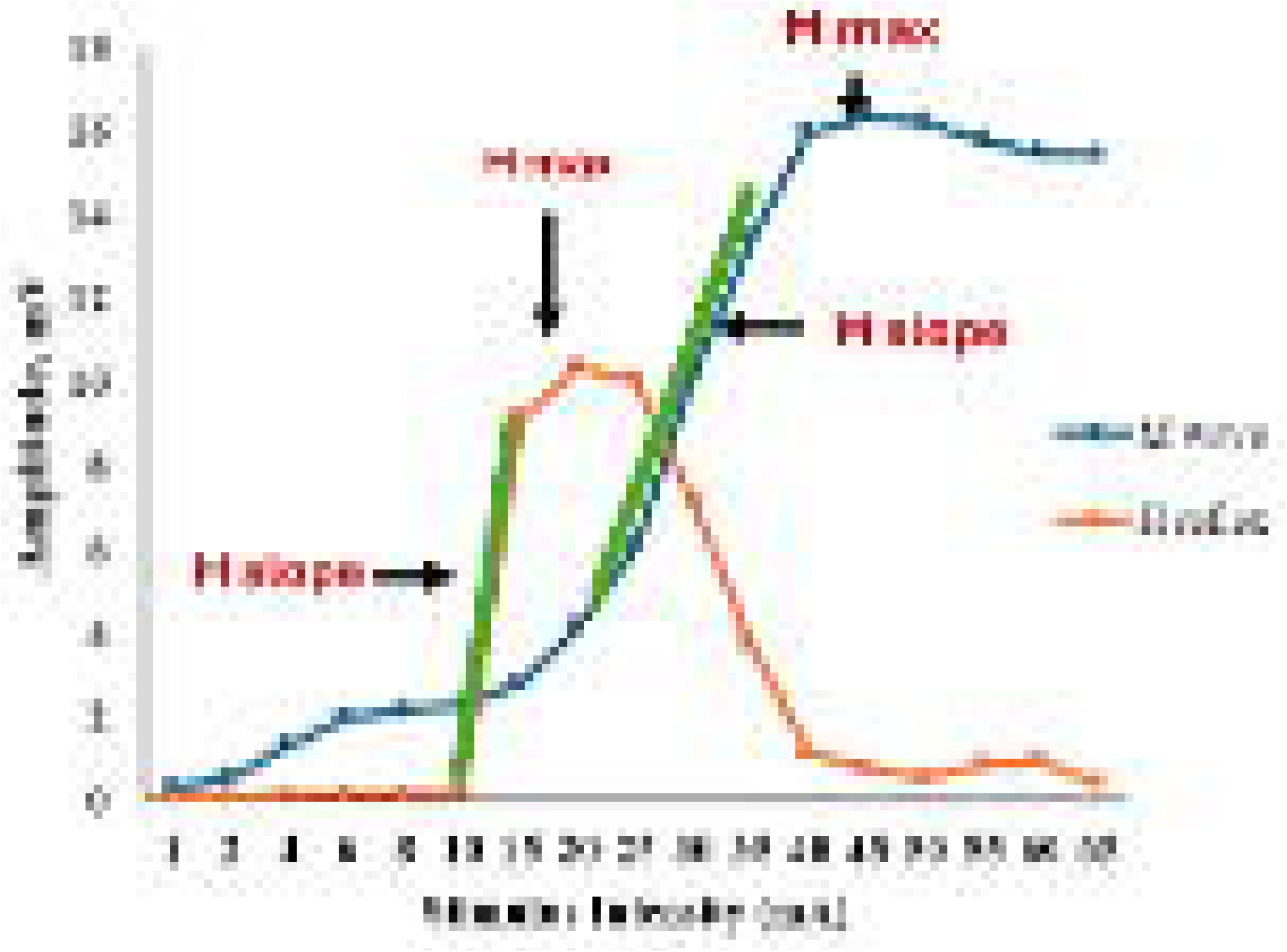

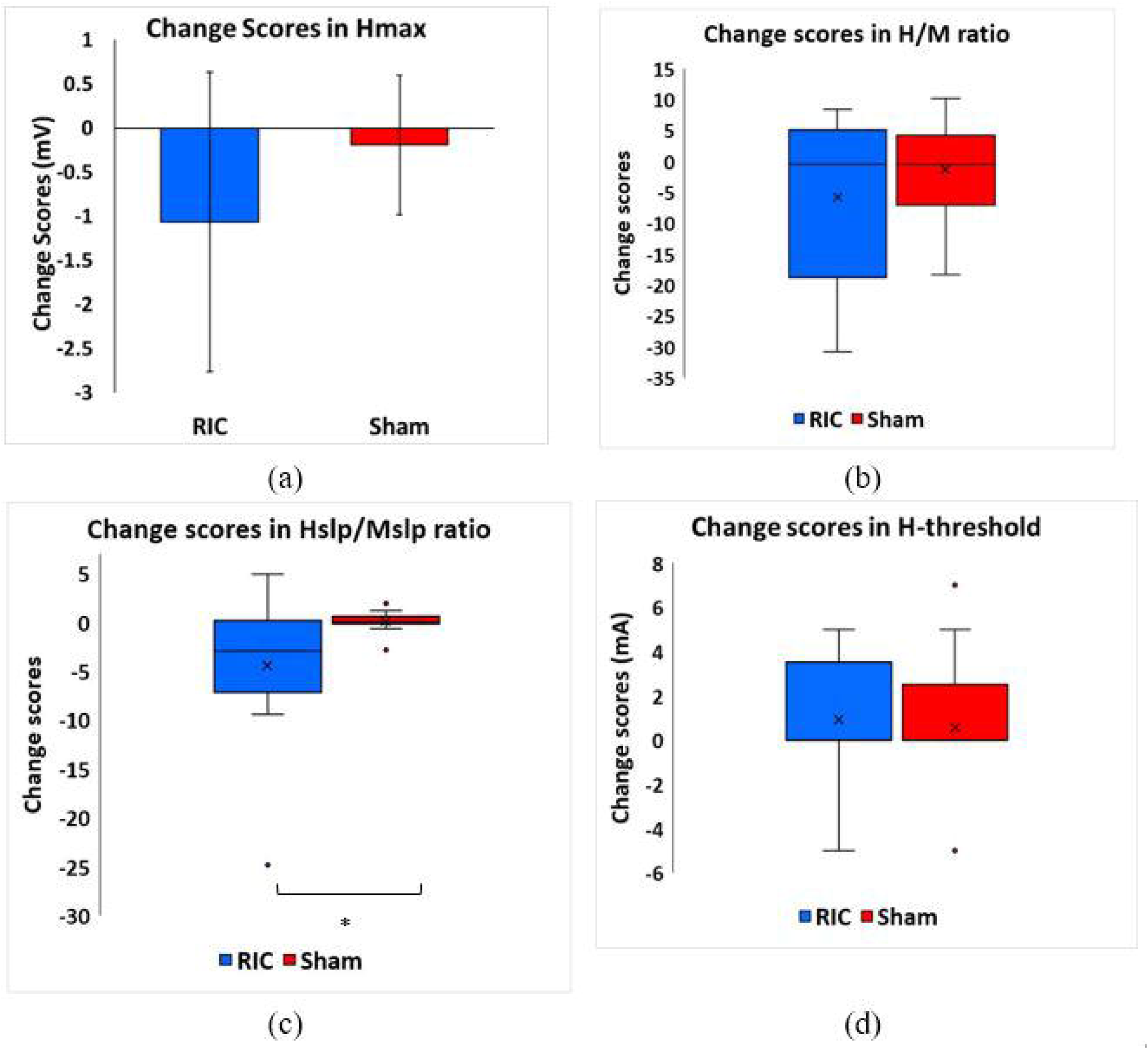

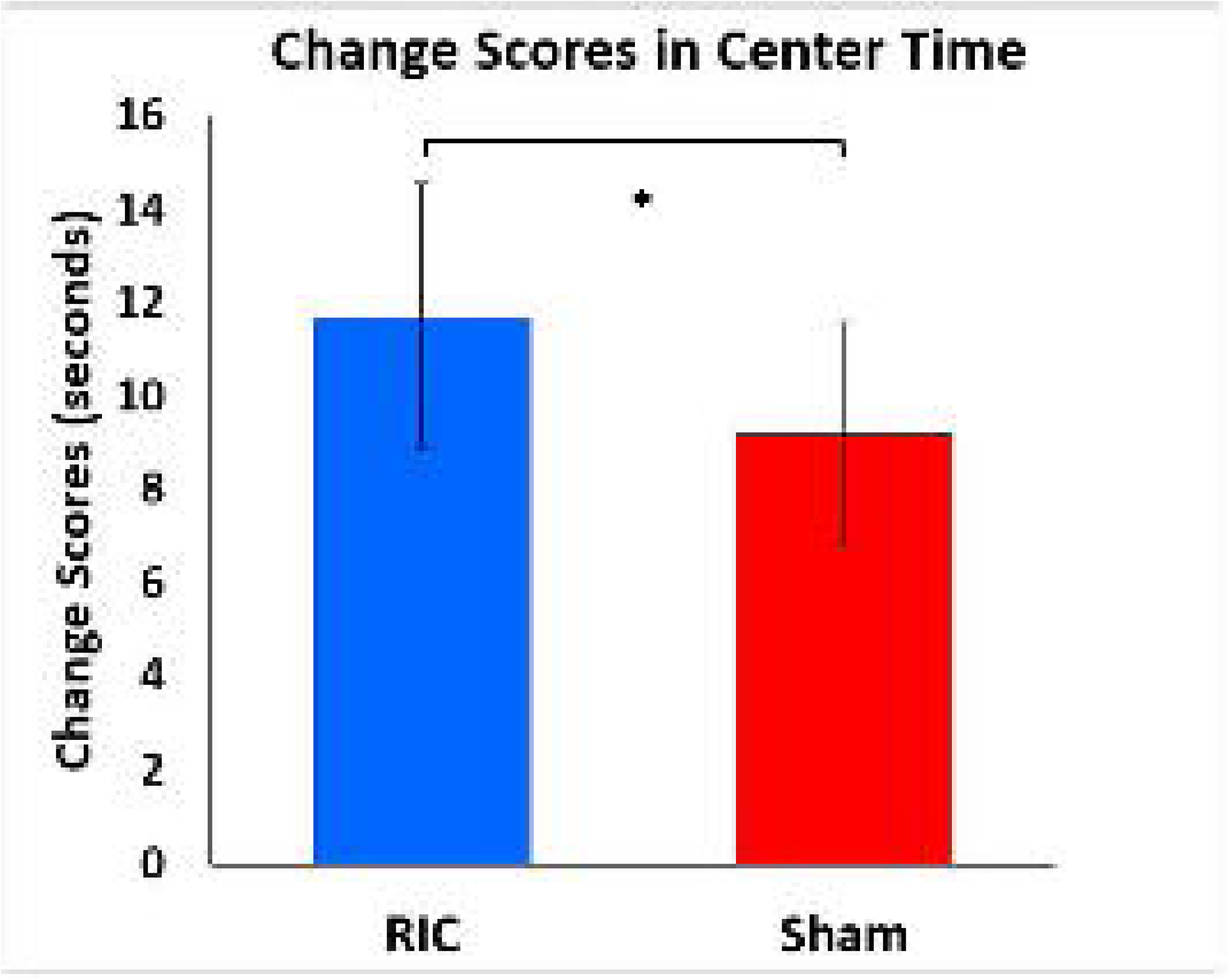

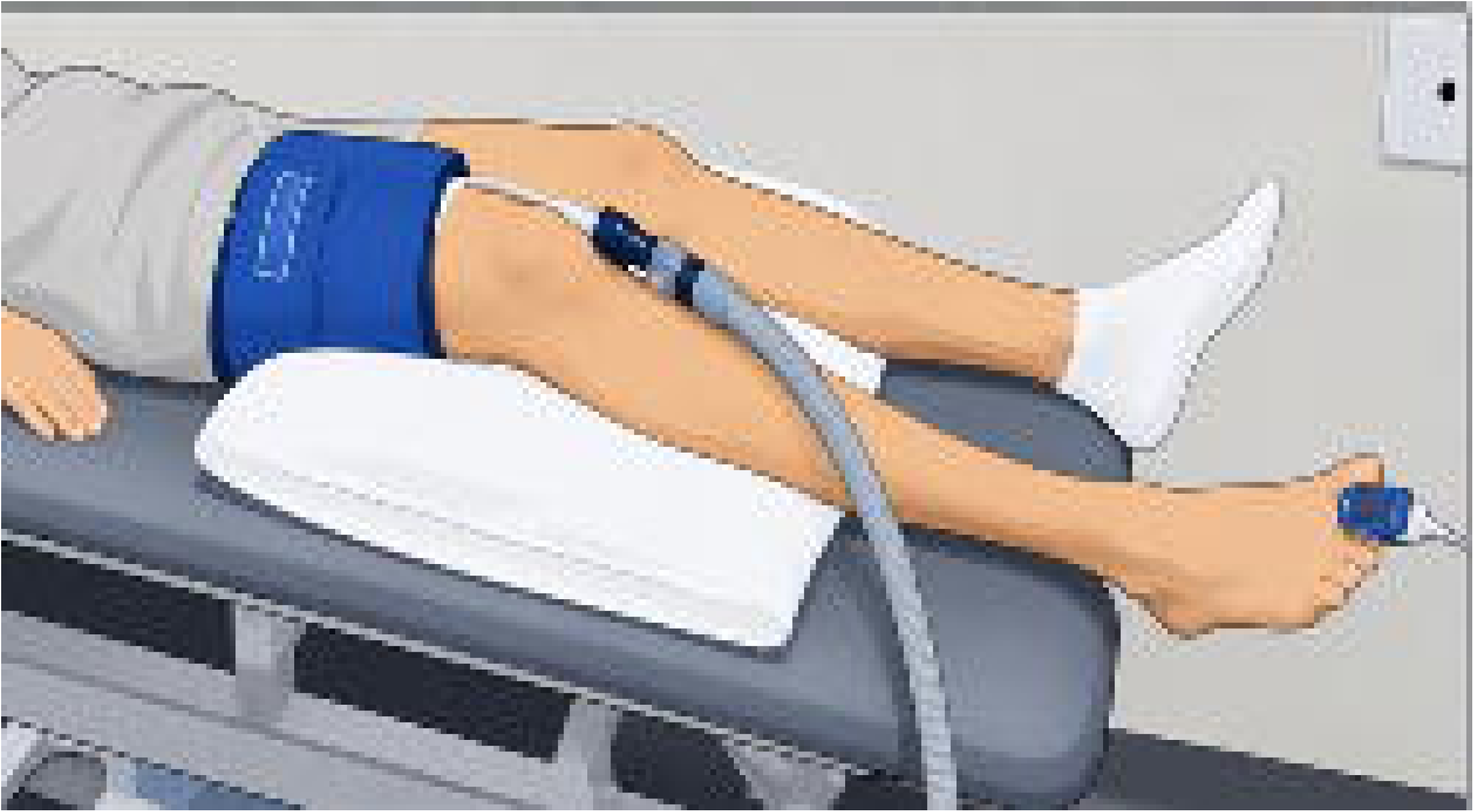

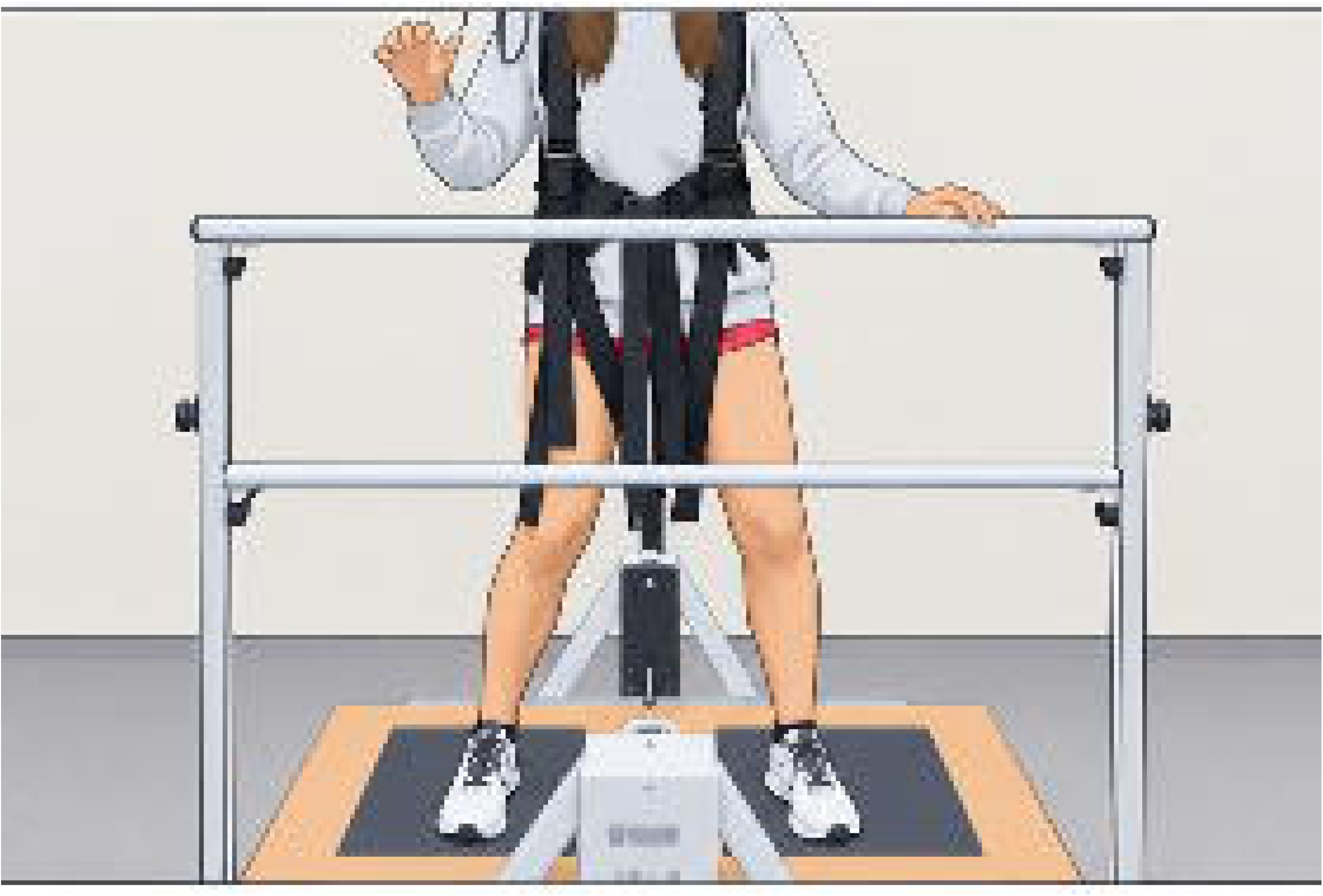

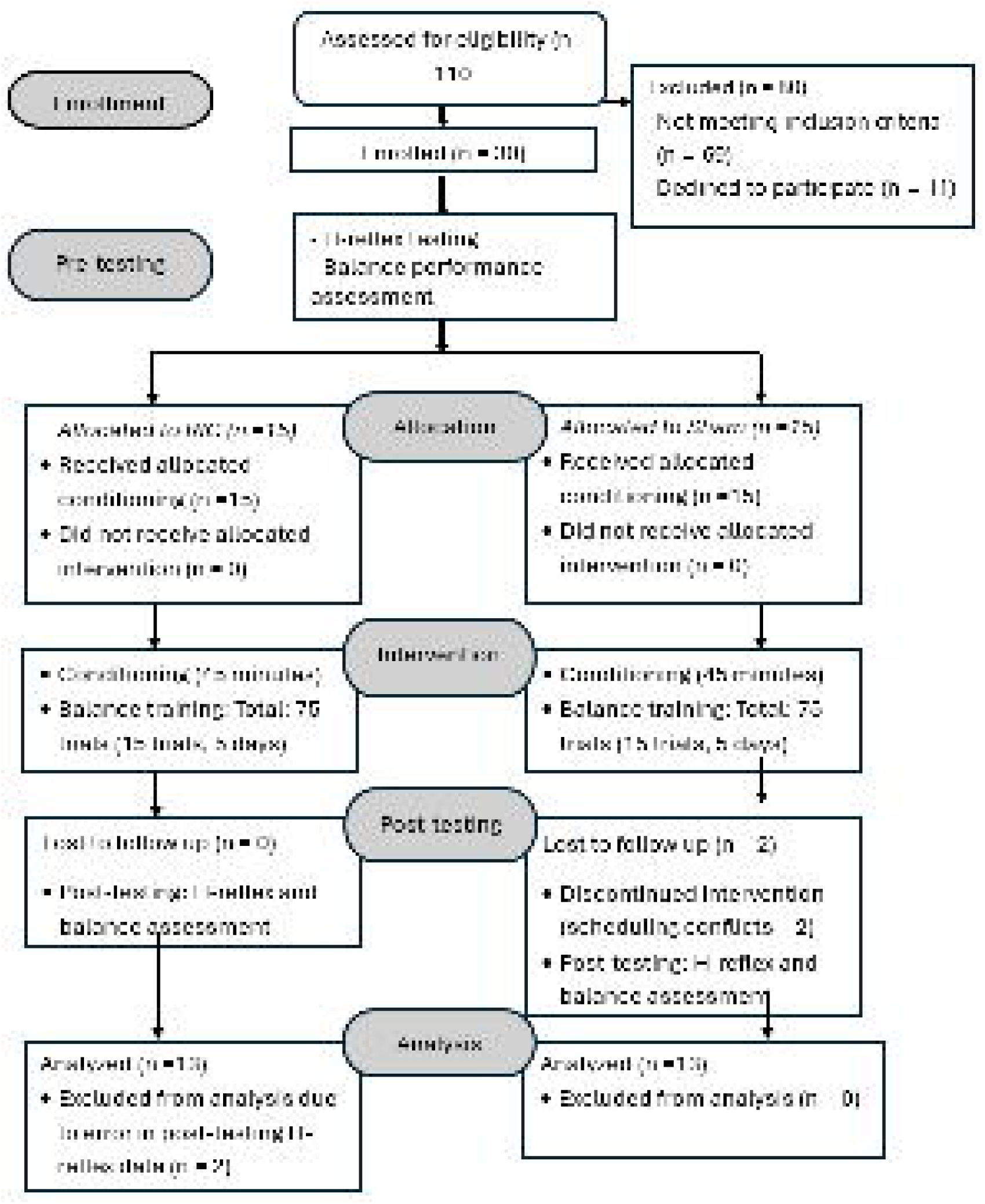

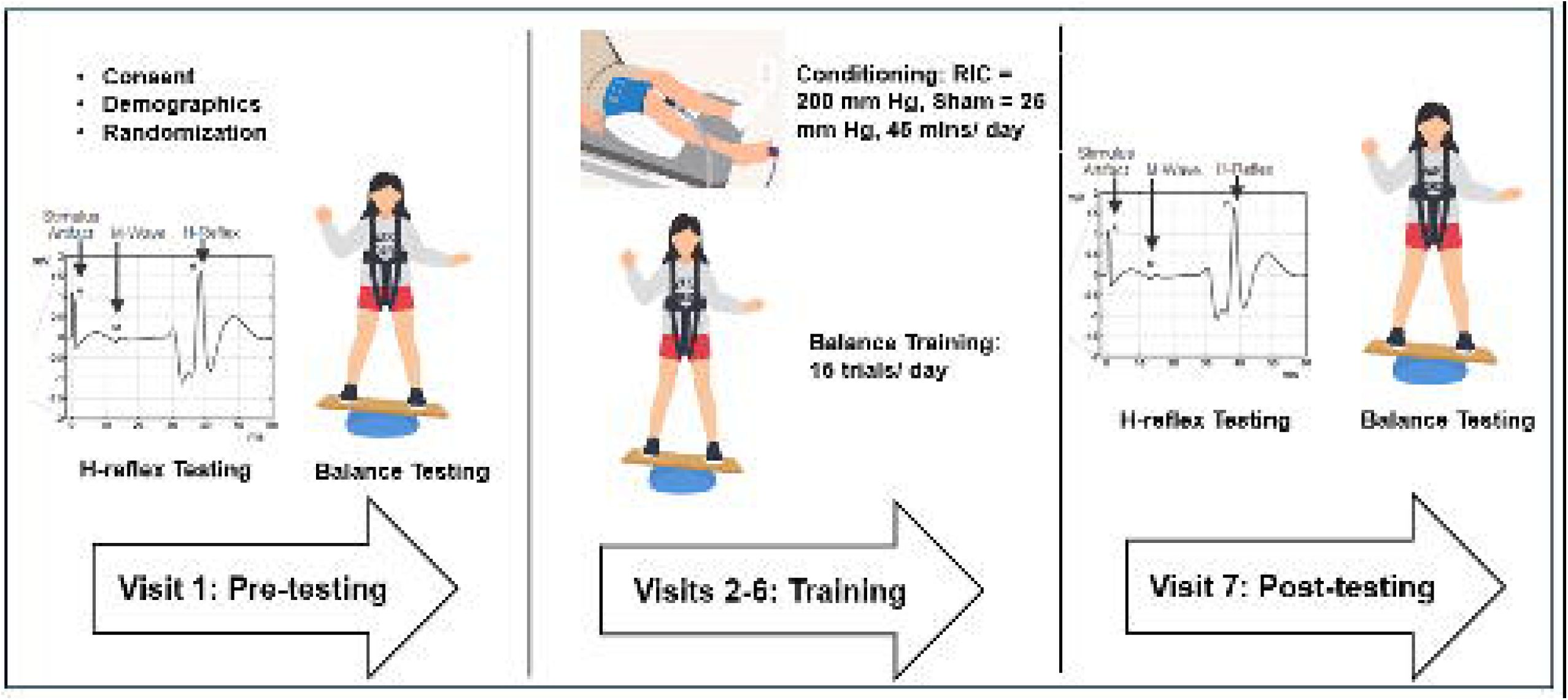

